# Impact on microbiology laboratory turnaround times following process improvements and total laboratory automation

**DOI:** 10.1101/2020.10.19.20213975

**Authors:** Carolyn Gonzalez-Ortiz, Alanna Emrick, Ying P. Tabak, Latha Vankeepuram, Stephen Kurtz, David Sellers, Megan Wimmer, Caitlin Asjes, Suhaireirene Suady Barake, Jacob Nichols, Fatma Levent

**Author notes:** **To whom correspondence should be addressed:** Fatma Levent, MD, Clinical Associate Professor, Pediatric Infectious Diseases / Department of Pediatrics, Texas Tech Health Sciences Center, Lubbock, TX. **Source of support:** This work was supported in part by Becton, Dickinson and Company.

## Abstract

**Introduction:** The impact of workflow changes and total laboratory automation (TLA) on microbiology culture processing time was evaluated in an academic-affiliated regional hospital.

**Materials and Methods:** A retrospective analysis of microbiological data in a research database was performed to compare turnaround time (TAT) for organism identification (ID) before and after implementation of TLA (2013 versus 2016, respectively). TAT was compared using the χ2 test for categorical variables and log-transformed t-test for continuous variables.

**Results:** A total of 9,351 predefined common and clinically important positive mono-bacterial culture results were included in the analysis. Shorter TAT (hours) in 2016 compared to 2013 (p<0.0001) for positive result pathogen ID were observed in specimen types including blood (51.2 vs. 70.6), urine (40.7 vs. 47.1), wound (39.6 vs. 60.2), respiratory (47.7 vs. 67), and all specimen types combined (43.3 vs. 56.8). Although shorter TATs were not observed from all specimen categories for negative result pathogen ID, TAT for all specimen types combined was shorter (p≤0.001) in 2016 compared to 2013 (94 vs. 101).

**Conclusions:** Total laboratory automation and workflow changes—including process standardization—facilitate shorter organism ID TAT across specimen sources.

## INTRODUCTION

Total laboratory automation (TLA) involves the integration of pre- and post-analytical processes through incorporation of multiple, modular systems to perform different tests, on diverse matrices, within the clinical microbiology laboratory, and typically consists of automation for plating, incubation, reading, and processing of specimens. Through reduced manual processing of specimens, standardization can be established to minimize the incidence of human error.1 Studies have demonstrated the clinical value of automating microbiological workflow including enhanced microbial growth, better colony isolation, reduced requirements for bacterial subculture, and reduced time to results.2–6 Previous work suggests that TLA can facilitate rapid reporting of microbiology culture testing, thereby informing the management of infectious disease and leading to enhanced patient care.6–8

This need for laboratory automation comes from an increased demand for clinical microbiology laboratory services, despite growing staffing challenges.9–11 Successful automation of a laboratory requires a thorough assessment and appropriate refinement of all laboratory workflow practices that might impact system efficiency.12, 13 This includes consideration of extending service delivery to a twenty-four-hour day, seven days per week (24/7) schedule.3, 13, 14 Such changes facilitate the maximum benefit from laboratory automation.

Previous work demonstrated that TLA can also improve laboratory workflow, efficiency, and reproducibility6, 9, 15, 16 while reducing errors.4, 8 Staffing reductions of approximately 30% have been reported after automation despite a 27% increase in average laboratory workload per day.3 Thus, TLA addresses the challenges associated with a high volume workload and allows managers to reallocate human resources to value-added activities.

In 2014, the University Medical Center (UMC), Lubbock, TX prioritized improvements to quality and efficiency in the microbiology laboratory, including moving to TLA. After an assessment of available TLA solutions, UMC leadership supported investment in the BD Kiestra™ TLA system (Kiestra; Becton, Dickinson and Company, BD Life Sciences—BD Integrated Diagnostic Systems, Sparks, MD, USA), which automates and standardizes sample setup, incubation, and reading. UMC became the second laboratory in the United States to implement Kiestra TLA and the first to both operate 24/7 as well as have all microbial sample types on the system. Preparation for the transition to automation included changes in laboratory policy and processes. The hypothesis here was that a combination of TLA and foundational workflow improvements (made in anticipation of the new technology associated with TLA) would result in a shorter post-TLA turnaround time (TAT). Although previous studies have evaluated the use of TLA for single specimen sources,5–7, 17–22 the goal here was to evaluate TAT across a wide range of specimen sources.

## MATERIALS AND METHODS

### Study Design

We compared microbiological result TAT before and after implementation of TLA and process improvements, including implementation of matrix-assisted laser desorption ionization time-of-flight mass spectrometry (MALDI-TOF) (Bruker Daltonik, Germany).23 The laboratory process improvement preparation started in 2014 and the TLA system went live in May 2015. The entire calendar years of 2013 and 2016 were used as pre-and post-periods, respectively, to evaluate TAT before and after laboratory technology and process changes. The year 2016 was selected as steady state to account for an implementation preparation period and a process-stabilization period after TLA implementation in 2015. This study was approved by the Quality Improvement Review Board at Texas Tech University Health Sciences Center. This work was conducted in a manner consistent with the ideals set forth by the World Medical Association Declaration of Helsinki regarding ethical conduct of research involving human subjects.

### Process improvement in preparation for TLA

#### Review of SOPs

Process improvements and total laboratory automation at the study site went through a stepwise and intricate process (see Figure 1). Initial steps were taken to ensure that process improvement evaluations were generated/updated that align with the College of American Pathologists guidelines for an accredited laboratory (https://www.cap.org/laboratory-improvement/accreditation/laboratory-accreditation-program). In April 2014, all SOPs were modified as needed according to procedural need; however, they remained aligned with American Society of Microbiology (ASM)24 and Clinical Laboratory and Standards Institute (CLSI) guidelines,25 as well as community practice standards, in an effort that involved networking with physician groups.

**Figure 1:**
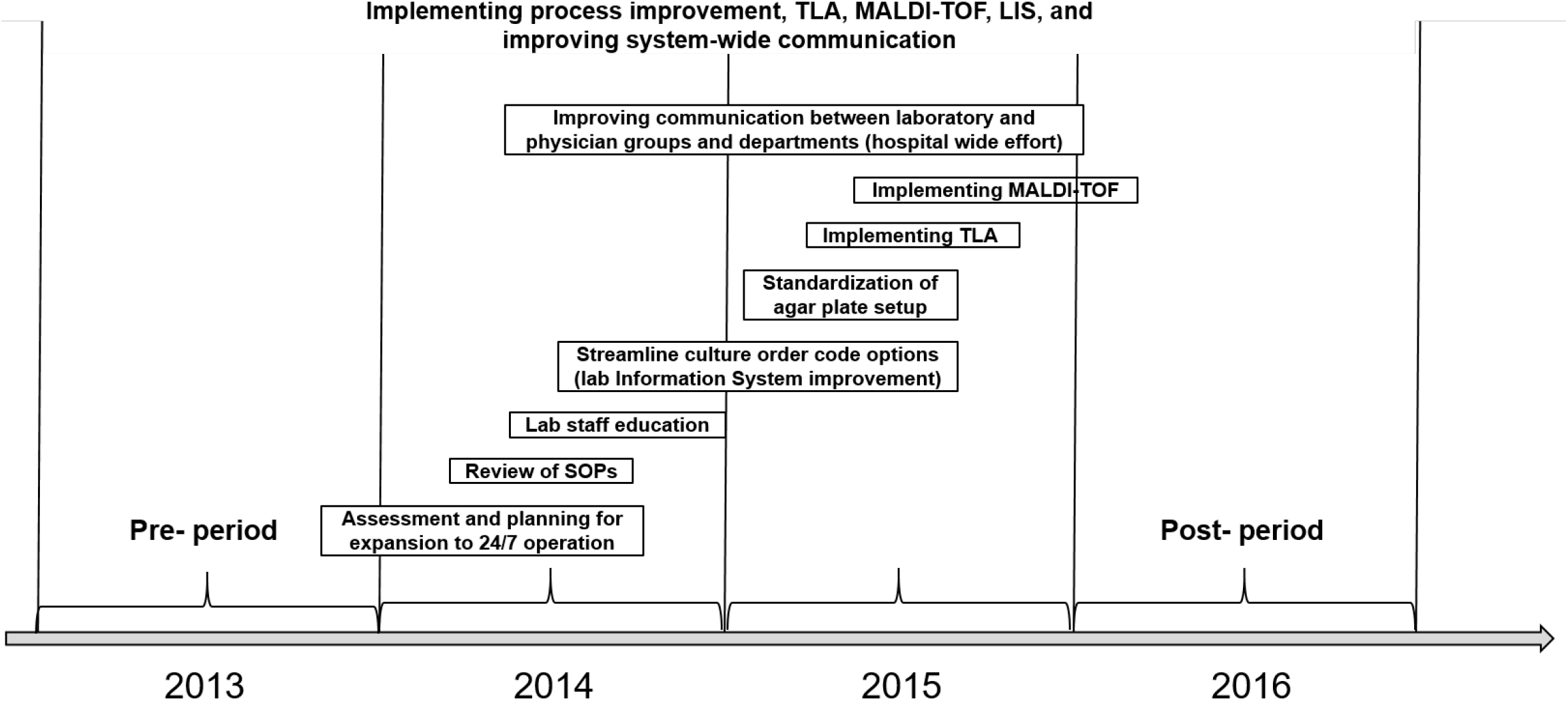
Microbiology Laboratory Quality Improvement Process Illustration. Kiestra TLA was implemented in May 2015. Abbreviations: TLA, Total laboratory automation; MALDI-TOF, Matrix-assisted laser desorption/ionization-time of flight mass spectrometry; LIS, Laboratory information system; SOP, Standard operating proceedure.

#### Staff education

All medical technologists (MTs) were actively engaged in professional education, assigned specific topics, and responsible for retraining the department members on their respective roles and monitoring compliance with the updated workflow practices. All MTs were educated and tested on best practice as described in the *Clinical Microbiology Procedures Handbook*.10

#### Streamlined culture order code options and standardization of agar plate setup

The UMC microbiology laboratory standardized culture setups and streamlined the number of order codes from 49 to 31. The standardization project resolved variability by narrowing plate setups to a single option and setting clear expectations for reading plates. Completing these projects simultaneously in November 2014 allowed for all MTs to be trained on all sources in preparation for the automation line, where cultures would be prepared for reading during setup. Digital imaging offered on the TLA system enabled plate images to be taken at ideal incubation times. This allowed reading of the plates to be optimally timed by reviewing the images rather than the plates themselves.14, 18, 26

#### Improved communication between laboratory and physician groups

Communication between laboratory management and physician/clinician groups during the TLA implementation period was approached here in a manner similar to that reported previously.27- 29 A diagnostic management team was created prior to commencing the transition to TLA. The team consisted of laboratory management and different physician/clinician groups, and included several goals. First, the team was created to ensure more consistent, open communication between the laboratory and clinicians. Initial meetings between physician/clinician and laboratory management included communication on process changes for ordering, testing, and downstream activities of reporting, charting, and consultation. Team meetings covered adjustments by the clinician to receive laboratory results in a reduced time frame in order to convert the time savings to a potential benefit for patients through evidence-based practices. Feedback from physician/clinician groups was received by the laboratory regarding TLA-related issues such as patient charting based on the previous collection day. Related to charting, discussion items included any changes in test selection and result interpretation based on TLA implementation. Laboratory updates were provided to physician/clinician groups at team meetings throughout the TLA implementation process. Post-implementation meetings revealed physician/clinician perspectives on progress based on initial goal setting for processes including reporting and charting. Feedback was also provided to the laboratory for continued improvement.

### Pre- and Post-TLA sample processing

During the 2013 study period, specimens arrived for processing throughout the day and specimen login to the laboratory information system was considered the initial time point for TAT (this also applies to 2016). For the conventional plating technique, urine specimens were streaked manually using calibrated loops onto blood and MacConkey agar plates and placed into incubators. Specimen plating was performed in batches and incubation initially occurred overnight (minimum of 18 hours). Plate reading was performed on four source benches, including blood/cerebral spinal fluid, wound/tissue/body fluid, urine, and stool/respiratory, each combining all plates into one batch during the morning. However, upon review, many plates were being read early at 16 hours per work day from 6 am to 10 pm (i.e. many plates that were prepared very early during the day shift [~6 am] were read at the end of the day shift [~10 pm]). Spot testing, which included oxidase catalase, pyrrolidonyl arylamidase, desoxycholate, indole, numerous latex agglutination tests, urease, and coagulase tests for Staphylococcus aureus, were conducted when required for identification. The BD Phoenix™ system (Becton, Dickinson and Company, BD Integrated Diagnostic Systems, Sparks, MD), A disk, P disk, RapID™ Systems (Thermo Fisher, Waltham, MA), or API® ID Strips (bioMérieux, Marcy-l’Étoile, France) were utilized for identification following all spot testing.

Kiestra TLA was implemented in May 2015 (Figure 1). Using TLA, specimens were inoculated upon reception (no batching) onto the appropriate agar plate type by the BD Kiestra InoqulA+ system (InoqulA), and plates were automatically transported via BD Kiestra ProceedA to ReadA Compact (incubation). Digital images of the plates were automatically acquired at 18, 42, and 66 hours depending on the sample type (i.e. 18 hours for day one, 42 hours for day two, and 66 hours for day three). Positive plates were sent to the ErgonomicA module, where a technologist performed MALDI-TOF MS for identification. Spot testing was reduced to only indole and desoxycholate for MALDI confirmation requirements, and coagulase and latex agglutination for a more rapid ID of S. aureus and β-hemolytic streptococci, respectively. Bruker MALDI-TOF MS was implemented in February 2016 for gram-negative IDs; gram-positive IDs were added in October 2016, and anaerobe and yeast IDs were added in April 2017. MALDI-TOF MS replaced identification by conventional methods, and times to result for positive specimens, preliminary negatives, and antimicrobial test reporting were recorded as described above. All other spot testing and ID methods were fully replaced with MALDI-TOF MS. A similar workflow was utilized to identify cultures as negative. Plates identified as no growth or no pathogens at 18 hours were further incubated for an additional 24 hours for a total of 42 hours before a second image analysis was performed. Times to result for positive specimens and preliminary negatives were as described above.

### Specimen Source and Organism Identification

Specimens were classified by source as blood, urine, wound, respiratory, intra-abdominal, and other. The top ten common and clinically relevant bacteria seen in the laboratory were Staphylococcus aureus, Escherichia coli, Klebsiella pneumoniae, Streptococcus pneumoniae, Streptococcus pyogenes (Group A), Streptococcus agalactiae (Group B), Pseudomonas aeruginosa, Proteus mirabilis, Enterococcus faecalis, and Enterococcus faecium. From the microbiology laboratory, results of 19.3% (2013) and 20.2% (2016) of the total results in each respective year are included in this study. The analysis was restricted to positive specimen results for mono-bacterial isolates of interest and negative culture results.7, 18, 24, 30

### Definition of definitive results TAT

We assessed definitive positive organism identification (ID) TAT as time from laboratory login of specimen to the earliest definitive positive result reporting time. Likewise, the TAT for negative results was from laboratory login to the earliest definitive negative result report.

### Data Source

De-identified digital microbiological data were captured in a research database (BD Insights) created by Becton, Dickinson and Company (Franklin Lakes, NJ).19, 20, 22 The dataset for the current study included specimen collection time, source, laboratory login time, and culture results reporting time.

### Statistical analysis

We analyzed organism ID reporting time over a 24-hour spectrum. Definitive positive organism ID and negative results TAT for pre-versus post-periods were compared. The χ2 test for categorical variables and log-transformed t-test for continuous variables were utilized for comparisons. A two-tailed p-value of <0.05 was considered statistically significant. All analyses were conducted using SAS version 9.4 (SAS Institute, Cary, NC).

## RESULTS

Overall 4,510 positive results were reported (Table 1). The mean TAT for all specimen types was significantly shorter after TLA and process change implementation (2106) compared to pre-TLA (2013) (all p<0.05).The overall TAT mean (standard deviation) was 43.3 (20.8) hours versus 56.8 (24.3) hours for years 2016 and 2013, respectively. Figure 2 depicts the comparison of cumulative distribution of TAT for positive IDs for the four most common specimen sources (blood, urine, wound, and respiratory; all p<0.05). For blood specimens, the ID TAT was 53% (2016) versus 17% (2013) by 48 hours from laboratory login time (Figure 2a). For samples derived from urine, wound, and respiratory specimens, a higher percentage of results was returned within 24 hours in 2016 compared to 2013. At least 40% of the urine, wound, and respiratory specimens had TAT <36 hours in 2016, whereas in 2013, 40% of wound and respiratory specimens had TAT >36 hours, and 40% of urine TAT exceeded 24 hours. For urine specimens, the ID TAT was 51% (2016) versus 14% (2013) by 36 hours from laboratory login time (Figure 2b). For respiratory specimens, the ID TAT was 59% (2016) versus 21% (2013) by 48 hours from laboratory login time (Figure 2c). For wound specimens, the ID TAT was 77% (2016) versus 39% (2013) by 48 hours from laboratory login time (Figure 2d).

**Table 1.** Descriptive statistics for definitive positive bacterial pathogen ID turnaround time (hours)

| Specimen | Year | n | Mean (SD) | Median (IQR) | p-value<sup>a</sup> |
| --- | --- | --- | --- | --- | --- |
| Blood | 2013 | 759 | 70.6 (28.9) | 62.8 (52.0, 74.5) | <.0001 |
|  | 2016 | 882 | 51.2 (19.2) | 46.6 (38.0, 59.1) |  |
| Urine | 2013 | 2,282 | 47.1 (14.9) | 43.6 (38.0, 48.9) | <.0001 |
|  | 2016 | 2,306 | 40.7 (18.8) | 35.5 (27.7, 47.9) |  |
| Wound | 2013 | 1,160 | 60.2 (25.6) | 58.6 (43.1, 69.2) | <.0001 |
|  | 2016 | 869 | 39.6 (21.3) | 32.8 (25.6, 47.0) |  |
| Respiratory | 2013 | 461 | 67.0 (23.5) | 65.3 (49.8, 74.7) | <.0001 |
|  | 2016 | 369 | 47.7 (24.9) | 41.9 (28.1, 57.7) |  |
| Intra-abdominal | 2013 | 23 | 69.9 (28.8) | 64.3 (53.1, 84.7) | 0.0368 |
|  | 2016 | 34 | 57.8 (32.1) | 49.9 (25.7, 78.2) |  |
| Other<sup>b</sup> | 2013 | 56 | 75.1 (57.4) | 64.5 (46.4, 89.8) | 0.0013 |
|  | 2016 | 50 | 52.6 (30.5) | 46.1 (29.3, 69.7) |  |
| Overall | 2013 | 4,741 | 56.8 (24.3) | 48.4 (41.1, 66.4) | <.0001 |
|  | 2016 | 4,510 | 43.3 (20.8) | 38.0 (28.7, 51.7) |  |

Abbreviations: SD, standard deviation; IQR, inter-quartile range (1<sup>st</sup>, 3<sup>rd</sup> quartile)
<sup>a</sup>Log-transformed t-test for continuous variables, utilized for comparisons of mean turnaround time.
<sup>b</sup>"Other" includes specimens from duodenal aspirate, ear, eye, quantitative tissue, quantitative bronchoscopy, environmental samples (surgical hardware, transfusion reactions), stool, cystic fibrosis cultures, dialysates, catheter tips, cerebral spinal fluid, genital, and throat.

**Figure 2:**
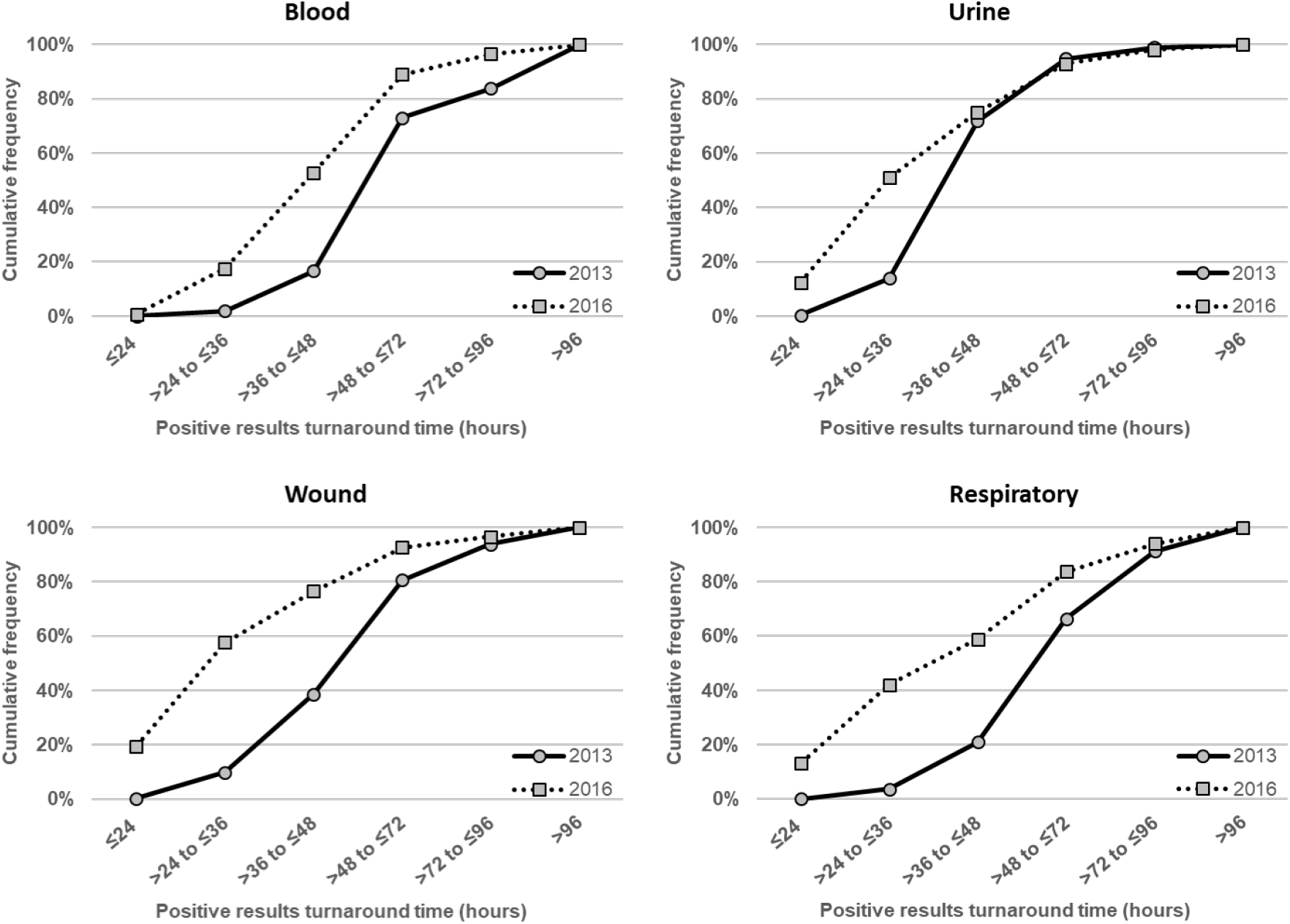
Comparison of cumulative distribution of definitive positive bacterial pathogen ID turnaround time following login by most common source: a) blood, b) urine, c) respiratory, and d) wound. All values p<0.05.

Overall 58,640 negative results were reported (Table 2). All sources except for respiratory and “other” showed shortened TAT (all p<0.05). Figure 3 depicts the comparison of cumulative distribution of negative result TAT for the four most common sources (blood, urine, wound, and respiratory; all p<0.05). For blood specimens, the TAT was 95% (2016) versus 73% (2013) by 128 hours from laboratory login time (Figure 3a). TAT for specimens derived from urine and wound showed the greatest reduction in 2016 compared to 2013. For urine specimens, the TAT was 83% (2016) versus 10% (2013) by 36 hours from laboratory login time (Figure 3b). For wound specimens, the ID TAT was 71% (2016) versus 36% (2013) by 96 hours from laboratory login time (Figure 3c). For respiratory specimens, the negative results TAT was 79% (2016) versus 86% (2013) by 96 hours from laboratory login time (Figure 3d). Negative results TAT was less variable across the time frame categories for blood and respiratory between 2013 and 2016. However, the 80th percentile of TAT results for blood were returned within 126 to 128 hours in 2016, while those in 2013 were delayed to between 128 to 240 hours.

**Table 2.** Descriptive statistics for negative result turnaround time (in hours)

| Specimen | Year | n | Mean (SD) | Median (IQR) | p-value<sup>a</sup> |
| --- | --- | --- | --- | --- | --- |
| Blood | 2013 | 16,393 | 131 (54) | 126 (122, 128) | <.0001 |
|  | 2016 | 17,905 | 125 (6) | 125 (123, 127) |  |
| Urine | 2013 | 8,115 | 44 (10) | 45 (40, 47) | <.0001 |
|  | 2016 | 8,880 | 30 (14) | 26 (22, 32) |  |
| Wound | 2013 | 1,100 | 108 (40) | 114 (74, 120) | <.0001 |
|  | 2016 | 957 | 82 (43) | 66 (50, 113) |  |
| Respiratory | 2013 | 1,204 | 62 (30) | 50 (45, 64) | <.0001<sup>b</sup> |
|  | 2016 | 1,460 | 71 (36) | 54 (47, 80) |  |
| Intra-abdominal | 2013 | 154 | 129 (26) | 117 (113, 156) | <.0001 |
|  | 2016 | 280 | 115 (40) | 122 (103, 132) |  |
| Other<sup>c</sup> | 2013 | 613 | 121 (15) | 118 (114, 123) | <.0001<sup>b</sup> |
|  | 2016 | 674 | 170 (27) | 175 (164, 184) |  |
| Overall | 2013 | 27,579 | 101 (58) | 121 (48, 127) | <.0001 |
|  | 2016 | 30,156 | 94 (47) | 123 (38, 126) |  |

Abbreviations: SD, standard deviation; IQR, inter-quartile range (1<sup>st</sup>, 3<sup>rd</sup> quartile)
<sup>a</sup>Log-transformed t-test for continuous variables, utilized for comparisons of mean turnaround time.
<sup>b</sup>For respiratory and "other" sources, the negative turnaround time was shorter in 2013.
<sup>c</sup>"Other" includes specimens from duodenal aspirate, ear, eye, quantitative tissue, quantitative bronchoscopy, environmental samples (surgical hardware, transfusion reactions), stool, cystic fibrosis cultures, dialysates, catheter tips, cerebral spinal fluid, genital, and throat.

**Figure 3:**
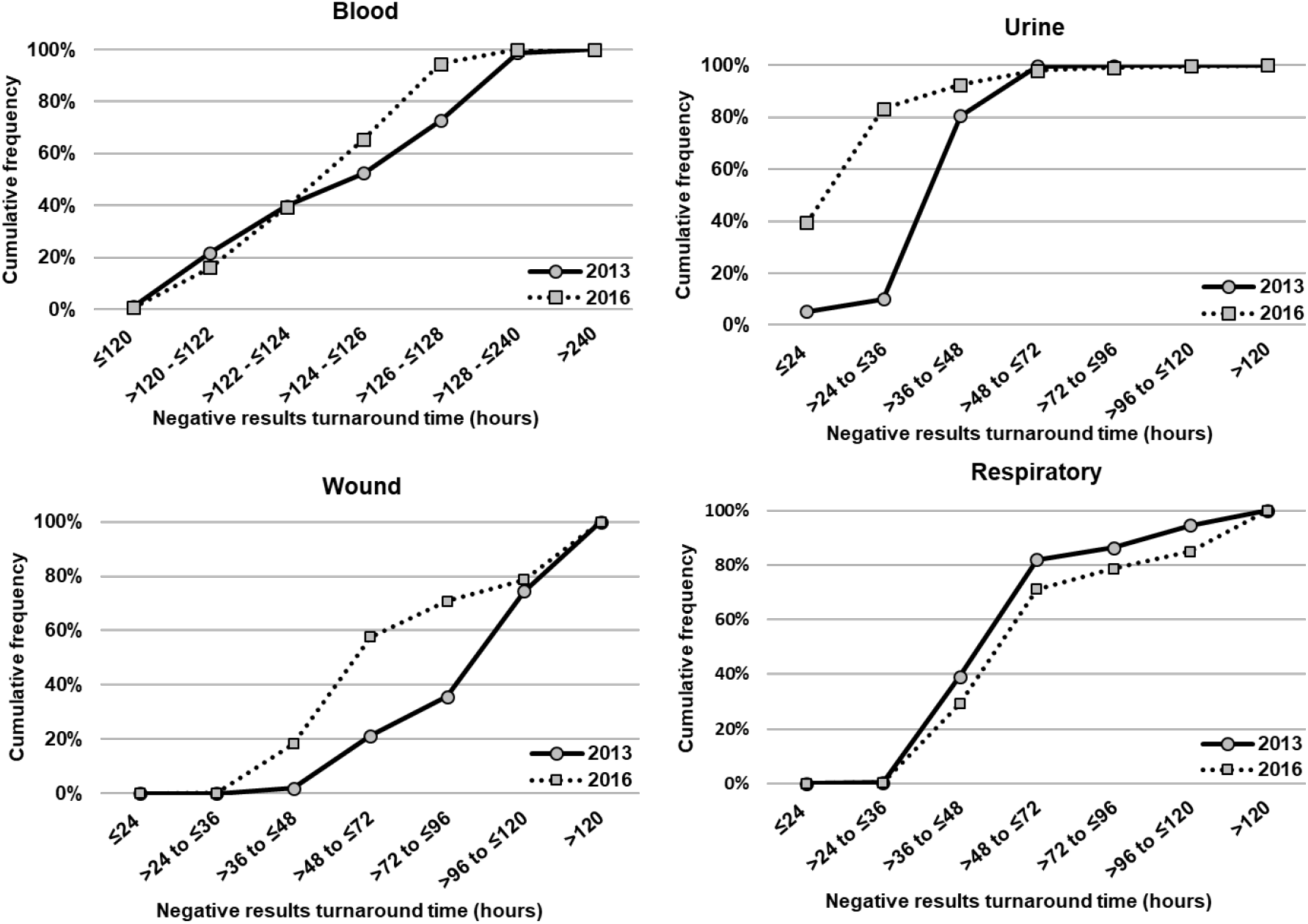
Comparison of cumulative distribution of definitive negative culture results turnaround time following login by most common source: a) blood, b) urine, c) respiratory, andd) wound. All values p<0.05. ^**a**^The extended time reported for blood cultures reflects an initial growth/detection phase prior to plating.

## DISCUSSION

The results here suggest that TLA, in conjunction with process changes that facilitate TLA, led to improvement in TAT for microbial ID. Although the observed shorter TAT was likely a joint effect of intricate laboratory process improvement and implementation of the TLA, these results are in agreement with previous studies that show a positive correlation between laboratory automation and faster TAT. 18–22, 28 In general, the overall laboratory process improvement interventions and TLA implementation (2016) led to shorter TAT for positive results compared to 2013. For the highest volume specimen, urine cultures, pathogen ID TAT improved by an average of 6.4 hours from 2013 to 2016. Since TLA processes and images the plates under consistent incubation conditions, facilitating more robust growth,18 a 16 to 30-hour growth period was no longer necessary, which likely facilitated earlier ID. Blood cultures showed significant improvement in TAT to pathogen positive ID by 19 hours, potentially enabling providers to update their course of treatment by almost an entire day for patients with bacteremia. In addition to urine and blood cultures, wound, respiratory, intra-abdominal, and “other” cultures showed improvements of 20.6, 19.3, 12.2, and 22.5 hours, respectively (Table 1, Figure 2). The use of ReadA Compact smart incubators, which reduce the incubation time needed, plus MALDI-TOF for ID of gram-negative organisms, which reduces the time to definitive ID, could contribute to improvement in TAT.

The implementation of laboratory process improvements incorporating TLA largely led to reductions in TAT for negative culture results. For urines, the average time to report a negative result in 2016 improved by 14 hours compared to 2013. Prior to TLA implementation, all negative urine cultures were read manually, which necessitated pulling plates from the incubator racks by hand. TLA standardizes incubation duration, so cultures are available to read at the appropriate time as opposed to times that might be dictated by staff availability or other daily workflow considerations. Digital imaging allows negative urines to be read on-screen, reducing the manual labor of removing each plate from the incubator, and occurred at the 18-hour mark with TLA. These approaches are consistent with current CLSI and ASM guidelines.24, 25 Most likely due to TLA capabilities including digital imaging and improved incubation times, the read time for the majority of urine cultures was reduced to one day, with the exception of suprapubic, straight catheter, cystoscopy, and nephrostomy sources. The results of negative wound cultures were obtained an entire day sooner. Process changes prior to TLA included the removal of thioglycolate broths from the wound cultures and a reduction in plate incubation times. Improvements in TAT for respiratory and “other” were not observed for negative results. The microbiology laboratory reported that respiratory and “other” specimens grew many isolates that were more difficult to identify at first pass. Analyses involved with this study were focused on single isolate cultures. Respiratory and “other” specimen types often grew isolates that were mixed, overgrown, and/or mucoid. Subcultures were often necessary, delaying ID TAT. For the mucoid specimens, repeated IDs were sometimes necessary, causing delays.

The significant time savings reported here were likely due to several factors. The overall laboratory process improvement interventions, including SOP update, staff education, and hospital-wide collaborations, are likely integral parts of the improvement in the shortened ID TAT. In addition, the TLA system aided in reducing TAT through a number of key technical components. Because ESwab™ (Copan Diagnostics Inc., Murrieta, CA, USA) and BD Vacutainer™ (Becton, Dickinson and Company, Franklin Lakes, NJ) system-collected urines could be fully automated, there was more than a 75% reduction in manual plate inoculation. The system includes a plate-streaking module, the InoqulA, which has been shown to improve colony isolation through automated plate streaking.2, 4, 5 BD Kiestra TLA also features a component called the ReadA Compact, a “smart” incubator, which does not require interruptions during the incubation process in order to improve growth conditions.7, 15 The physical connection between the InoqulA module, ReadA Compact, and the ProceedA (a track that automates movement of specimens between modules) minimizes the time that plates spend in the open air at room temperature; plates are instead transported by the ProceedA to the ReadA Compact within minutes. Moreno-Camacho et al demonstrated that the ReadA Compact uses enhanced organism growth cycles and improved the rate of viable organisms available for further identification and accurate susceptibility testing.15 Standardization of specimen preparation and detection methods has been shown to limit the potential for variability and to reduce manual reconciliation and interpretation by individual technologists.4

Previously, cultures received a non-standardized 24 hours of incubation. Incubation times varied from 16 to 30 hours of first incubation prior to TLA and process changes. Ongoing process review and refinement to maintain adherence to current guidelines also precipitated further removal of redundant or obsolete steps from manual processes (such as certain broth incubations) for specific specimen types. Interestingly, we found that laboratory process improvement and TLA implementation led to a more even distribution of clinical results during a 24-hour time spectrum (Figure S1). However, in line with previous discussion regarding work distribution following TLA implementation,31 we view a more even distribution of workflow across a 24-hour work period as an indicator of improved workflow efficiency that reflects reductions in unnecessary or excessive work and/or idle/waiting time. Automation of negative result identification further allows laboratory personnel to focus on other important aspects of clinical work, for example by allowing more time to read positive plates that require technical skill. This idea is supported by previous work, as Greub et al estimated that 50–70% of the time spent by full-time technicians may be saved by automation of pre-analytical sample preparation and plate inoculation alone.32

This study has limitations. Since this is a retrospective observational study of a microbiology laboratory TAT improvement in a real-world setting, each intervention was not pre-designed to last long enough to allow for assessment of independent impact. The standardization and workflow improvement activities were integral parts of TLA implementation, in which each intervention often overlapped with other interventions. Thus, the impact of each component attributable to the shortened TAT could not be measured independently. This resulted in an inability to clearly delineate the proportion of TAT improvement attributable to process improvement versus TLA. Likewise, MALDI-TOF was not fully integrated (to include yeast identification along with Gram negative and Gram positive bacteria) until 2017, which admittedly affected our capability to fully assess the impact of TLA at the time of study. A goal for future studies will be to further investigate this continued quality improvement. Another limitation is that we did not directly assess whether improved (shorter) TAT was associated with improved patient outcomes, which is beyond the scope of this study. Nevertheless, performance values following TLA using Kiestra have previously been demonstrated.17, 26, 33 Furthermore, previous work has been able to provide more direct evidence for a benefit of specific automation components after the implementation TLA. For example, Theparee et al 20 were able to isolate the impact of TLA (median ID TAT decrease of ~2 hours compared to pre-TLA) and of MALDI-TOF (median ID TAT decrease of ~3 hours compared to TLA alone).20 Mutters et al combined TLA and MALDI-TOF together versus conventional testing. However, they completely implemented the MALDI-TOF library at one time, as opposed to the stepwise implementation of Gram negative, Gram positive, and yeast organisms over the course of one to two years as occurred at this study site. In that study TLA combined with MALDI-TOF led to an antibiotic switch in 12% of cases (in addition to those cases in which a switch was based on Gram staining) as well as shortening the first 24-hour incubation time. Ultimately, however, randomized, controlled trials will be required to better “…evaluate the actual benefits of an automated system in terms of morbidity, mortality, and health care costs.”6

### Conclusions

In contrast to previous studies on the impact of TLA on clinical processes involving a single specimen type,7, 18, 19 this study describes the significant impact of TLA and related process changes on decreased TAT in microbial ID across a broad range of specimen types. A combination of TLA and associated workflow improvements contributes to shorter definitive organism ID TAT across all specimen sources and shorter TAT for negative results for most specimen sources. Overall, there was an average of approximately 13.5 hours improvement in TAT to organism ID across all subsets of cultures for which data were analyzed. Additionally, the overall TAT to negative result for all cultures, even with the regression of some cultures due to process change, improved an average of approximately 7 hours. Future studies could further investigate the relationship of operationalizing TLA technology and optimizing workflow, and potentially identify positive impacts on patient care by informing and thereby enabling physicians to make antibiotic therapy choices sooner.

## Data Availability

Data requests should be directed to David Sellers at David.Sellers[at]bd.com

## ACKNOWLEDGEMENTS

The authors thank Devin S. Gary, PhD (Becton, Dickinson and Company, BD Life Sciences – Diagnostic Systems) and Andrea Gwosdow, PhD (Consultant for Becton, Dickinson and Company, BD Life Sciences – Diagnostic Systems), for insight, discussion, and review during the preparation of this manuscript. The individuals acknowledged here have no additional funding or additional compensation to disclose.

## DISCLOSURES

This study was approved by the Quality Improvement Review Board of the study site. The researchers at Texas Tech University Health Sciences Center, University Medical Center, Lubbock, TX, and Becton, Dickinson and Co., Franklin Lakes, NJ conducted the analysis as their normal working responsibilities. YPT, LV, SK, DS, MW, and CA are current or former employees of Becton, Dickinson, and Company, the manufacturer of the BD Kiestra TLA system implemented by the study site. AE, CGO, SS, JN, and FL do not have any other disclosures. All authors provided final approval of the manuscript and agree to be accountable for the accuracy and integrity of this work.

## Abbreviations

TLA: total laboratory automation;
SOP: standard operating procedure;
MALDI-TOF: matrix-assisted laser desorption ionization time-of-flight mass spectrometry;
TAT: turnaround time;
ID: identification;
ASM: American Society of Microbiology;
CLSI: Clinical Laboratory Standards Institute;
MT: medical technologist

## Figure Legends

**Supplemental Figure 1:**
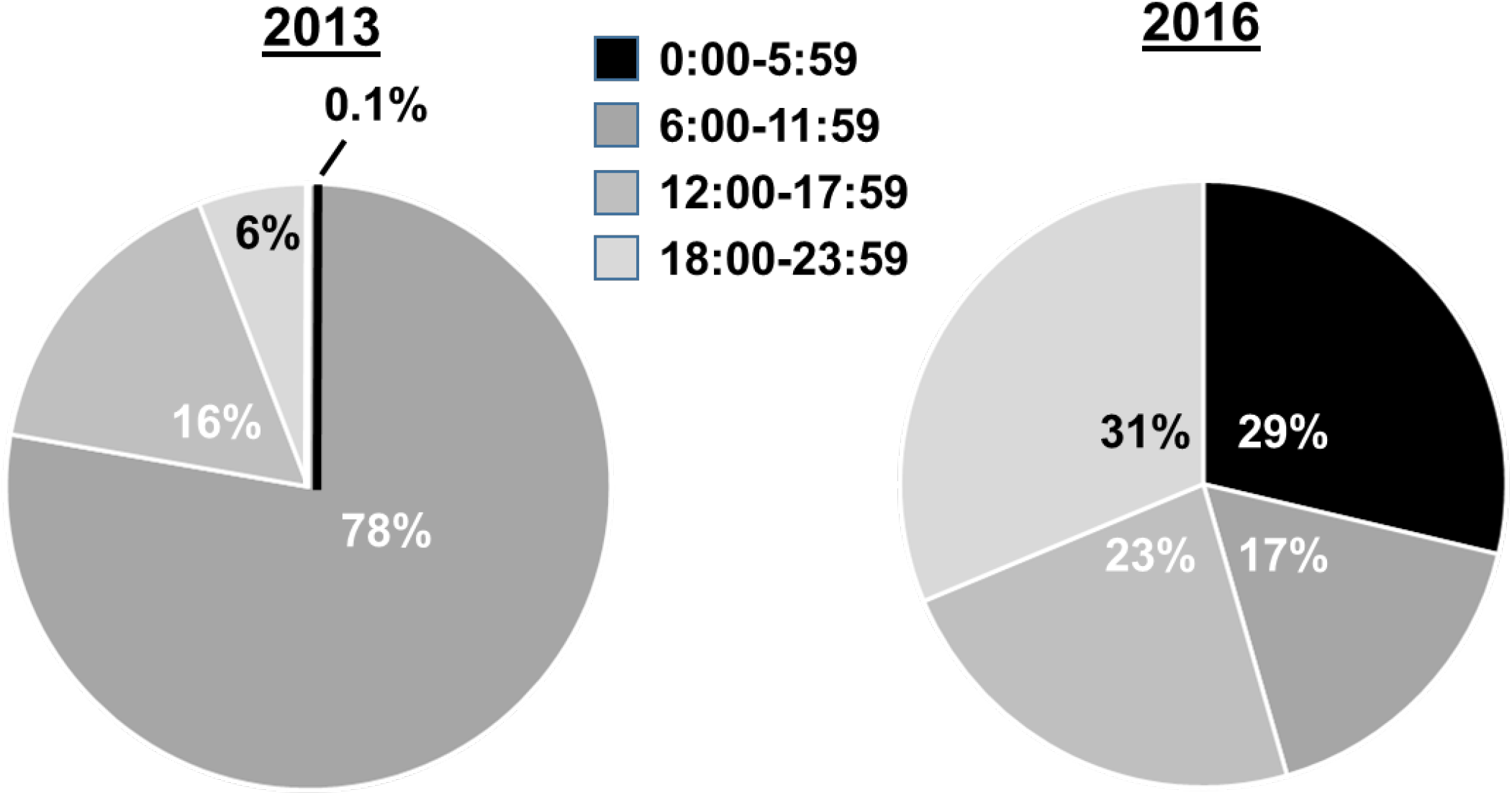
Organism ID reporting time distribution during a 24-hour spectrum.

## Notes

### Clinical Trial

This was not a clinical trial.

### Author Declarations

Texas Tech University Health Sciences Center - Institutional Quality Improvement Review Approval number 17005; expiration date: 05/15/2021.

### Summary of Updates

Abstract (original), lines 12-13: "were observed in specimen types including blood (40.7 vs. 47.1), urine (30 vs 44), wound (39.6 vs. 60.2), respiratory (47.7 vs. 67), and all specimen types, combined (43.3 vs. 56.8). Although shorter TAT were not observed from all" Changed to: "were observed in specimen types including blood (51.2 vs. 70.6), urine (40.7 vs. 47.1), wound (39.6 vs. 60.2), respiratory (47.7 vs. 67), and all specimen types combined (43.3 vs. 56.8). Although shorter TATs were not observed from all

